# Clustering of Social Determinants of Health and their Association with Adverse Cardiovascular Outcomes in Atrial Fibrillation

**DOI:** 10.1101/2025.11.20.25340708

**Authors:** Yusheng Zhou, Jonathan Houle, Valeria Raparelli, Colleen M. Norris, Louise Pilote

## Abstract

**Background:** Despite improvements in diagnostic and therapeutic options, individuals with atrial fibrillation (AF) remain at high risk for major adverse cardiovascular events (MACE). Social determinants of health (SDOH) are strongly associated with cardiovascular outcomes, yet the complex patterns through which these factors cluster to create distinct vulnerability profiles remain poorly understood.

**Methods:** We conducted a latent class analysis utilizing data from the UK Biobank cohort, examining 15 SDOH indicators across economic, psychological, and neighborhood domains among 3,842 participants with AF (35.1% female). Multi-group latent class analysis (LCA) evaluated differences in vulnerability pattern distribution, and explored whether SDOH clusters of vulnerability varies by sex. Cox proportional hazards models assessed the association between identified SDOH clusters and composite cardiovascular outcomes comprising major adverse cardiovascular events (MACE) and all-cause mortality.

**Results:** We identified five distinct SDOH vulnerability clusters based on multi-group LCA: 1) low vulnerability across all domains; 2) primarily economic vulnerability; 3) primarily neighborhood-related vulnerability; 4) economic and neighborhood vulnerability with favorable psychological conditions; and 5) high vulnerability across all domains. Male participants demonstrated a higher representation in more advantaged profiles than their female counterpart (Cluster 1: 37.0% vs. 26.6%; Cluster 4: 25.3% vs. 14.1%). Female participants exhibited the greater representation in the overall highest vulnerability cluster(Cluster 5: 35.1% vs. 25.2%) as compared with male participants. Compared to Cluster 1, individuals with AF from classes with adverse economic conditions (Cluster 2, 4 and 5) had a higher risk of MACE events with individuals in Cluster 5 having twice the risk. No sex interactions were observed with SDOH clusters in their association with MACE.

**Conclusions:** We identified five distinct SDOH vulnerability patterns among individuals with AF, revealing economic determinants as pivotal drivers of cardiovascular risk. These findings provide an evidence-based framework for implementing precision medicine approaches that incorporate comprehensive SDOH assessment into AF clinical management.

## Introduction

Despite widespread use of anticoagulation therapy and improvement in both screening and treatment strategies, individuals with atrial fibrillation (AF) (1) still experience high rates of major adverse cardiovascular events (MACE) such as stroke, transient ischemic attacks, arterial thromboembolic events, myocardial infarctions and cardiovascular mortality (2–4). While traditional clinical risk factors for MACE in participants with AF are well established, growing evidence suggests that social determinants of health (SDOH) play a crucial role in cardiovascular outcomes (5,6). SDOH, defined as the conditions in which people are born, grow, live, work, and age (7,8), have been shown to influence various aspects of AF care, including treatment adherence, healthcare utilization, and clinical outcomes (9).

While there is overwhelming evidence supporting SDOH determinants (e.g., income, education, social support) as upstream drivers (10–13), less attention has been paid to how SDOH intersect to create distinct patterns of vulnerability. In fact, SDOH factors rarely manifest in isolation. Instead, they tend to demonstrate co-occurrence patterns and potentially interact in ways that may amplify their impact on health outcomes (14,15). This phenomenon is particularly relevant for individuals with chronic disease, who frequently face multiple social and economic challenges that may influence their capacity for effective disease self-management and healthcare engagement (5,16).

In the present study, we utilized multi-group latent class analysis (LCA) models (17) to identify distinct patterns of SDOH vulnerability among individuals with AF and examine their associations with MACE outcomes and sex differences. This method is particularly relevant for studying the impact of SDOH, as they tend to cluster and interact in complex ways that may not be apparent when examined individually. This approach is therefore useful for revealing underrecognized mechanisms that might be targeted with precision-based social intervention strategies for reducing cardiovascular risk.

## Method

### Source and Study Population

Data were analyzed for the UK Biobank, a large-scale prospective cohort study of approximately 500,000 participants aged 40-69 years, recruited between 2006-2010 across the United Kingdom (18). Participants were evaluated at dedicated centers across the UK, where both self-reported and electronic health record data were collected, and were subsequently monitored during follow-up. (19). For this analysis, participants with a diagnosis of AF were identified through hospital admission records (International Classification of Disease 10 (ICD-10) code I48) and self-reported medical history at baseline. To maintain temporal proximity between SDOH assessments at enrollment and AF diagnosis, we restricted inclusion to participants diagnosed with AF within two years before or after enrollment. The UK Biobank study received approval from the Northwest Multi-Centre Research Ethics Committee (11/NW/0382) (https://www.ukbiobank.ac.uk/learn-more-about-uk-biobank/about-us/ethics). Participants provided informed consent at baseline. Our analyses were conducted under the UK Biobank application number 45551.

### Definition of SDOH variables

SDOH variables selected for the LCA to identify clusters were based on availability in the UK biobank database (20,21). Fifteen SDOH were included and categorized across three domains: i) economic factors including household income, highest education attained, employment status (employed vs unemployed or retired) and accommodation ownership; ii) psychological factors (i.e. living alone, social isolation, social support, emotional distress, and social inactivity; iii) neighborhood characteristics (i.e. area education quality level, material deprivation, housing quality, crime rate, greenspace remoteness, and blue space remoteness (Table S1). Each SDOH was coded using a standardized approach where 1 represents unfavorable social conditions and 0 indicates favorable circumstances. For missing values, LCA applies full-information maximum likelihood estimation, which derives parameter estimates while retaining all available data (22).

### Covariates

Baseline demographic, clinical and lifestyle variables were included as covariates. Demographic characteristics in this study refer to age, sex and self-reported race (23–25). Clinical factors included a history of heart failure, hypertension, diabetes, chronic kidney disease stages 3 or more, chronic obstructive pulmonary disease (COPD), anticoagulant use, body mass index (BMI). In addition, lifestyle variables included smoking history and alcohol consumption.

### Outcomes

The primary outcome was a composite of first occurrence of MACE (i.e., non-fatal myocardial infarction, non-fatal stroke, and cardiovascular death) and all-cause mortality. Secondary outcomes included the individual components of MACE, and non-CV mortality, separately. Time-to-event was defined as the interval between AF diagnosis and the first occurrence of the outcome. To ensure appropriate temporal sequencing and minimize immortal time bias, participants who experienced outcome events at or before the AF diagnose were excluded. Participants were followed from the date of AF diagnosis until the first occurrence of MACE, death, loss to follow-up, or the study end date (December 31, 2022).

### Statistical analysis

Latent class analysis (LCA) was employed to identify distinct patterns of SDOH vulnerability among AF participants (17). Observed SDOH indicators were used to determine the most meaningful clusters (26). Rather than generating phenotypes, LCA identifies homogeneous subgroups (27). Given established evidence demonstrating significant sex-based variations in SDOH distributions, health behaviors, and cardiovascular risk profiles (29–31), we employed multi-group LCA with self-reported biological sex as the grouping variable.

Multi-group LCA with sex as the grouping variable examines sex-specific distributional differences while testing measurement invariance across groups. Model selection incorporated multiple statistical criteria including Bayesian Information Criterion, with optimal class enumeration determined through systematic evaluation of 2-6 class solutions. Detailed LCA methodology, including model selection criteria, measurement invariance testing procedures, and technical specifications are provided in the Supplementary Methods (Supplement method). Cox proportional hazards regression models evaluated associations between identified vulnerability clusters and the composite CV outcomes. Models were adjusted for covariates. Proportional hazards assumptions were verified using scaled Schoenfeld residuals.

Statistical analyses were conducted using RStudio, poLCA statistical software package version 1.6.0.1(28), glca statistical software package version 1.4.2(29). Statistical significance was defined as p<0.05, with results presented as hazard ratios and 95% confidence intervals.

## Results

### Study Population Characteristics

The study cohort comprised of 3,842 participants with AF, of whom 35.1% were females, with a mean age 62.5 years (SD = 6.1), and an average follow-up duration of 10.1 years (SD = 3.8). A higher proportion of males experienced the composite outcome (29.1%) compared with females (21.3%). Males had a higher prevalence of diabetes, while no differences were found in other health conditions (Table 1). Lifestyle behaviors also differed by sex, with males reporting higher rates of smoking and alcohol consumption.

**Table 1.** Percent distributions of characteristics and outcomes of participants by latent class.

| **Variables** | **Cluster1**<br>**(N=972)** | **Cluster2**<br>**(N=1023)** | **Cluster3**<br>**(N=475)** | **Cluster4**<br>**(N=831)** | **Cluster5**<br>**(N=541)** |
| --- | --- | --- | --- | --- | --- |
| **Demographics**<br>**(%)** |  |  |  |  |  |
| Age (≥ 65 years) | 30 | 59.1 | 28.8 | 55.2 | 42.5 |
| Male | 72.5 | 58.6 | 69.5 | 62.5 | 62.3 |
| Race (Not white) | 1.6 | 1.8 | 7.2 | 2.9 | 6.5 |
| **Comorbidities**<br>**(%)** |  |  |  |  |  |
| Heart Failure | 2 | 2.4 | 4 | 3.2 | 5 |
| Hypertension | 17.5 | 28.4 | 19.6 | 30.2 | 33.8 |
| Vascular diseases | 1.9 | 3.2 | 1.9 | 2.8 | 6.1 |
| Diabetes | 3.9 | 5.5 | 6.3 | 6.5 | 9.1 |
| COPD | 1.6 | 4.7 | 3.4 | 6.7 | 14 |
| CKD | 1.9 | 3.7 | 2.1 | 4.3 | 5.2 |
| BMI | 28.4 ± 4.8 | 28.5 ± 5.1 | 28.6 ± 5.4 | 29.5 ± 5.6 | 30.1 ± 6.1 |
| **Lifestyle**<br>**behavior (%)** |  |  |  |  |  |
| Smoking | 51 | 50.5 | 52.2 | 57.2 | 66 |
| Alcohol<br>consumption | 59.9 | 47.2 | 51.2 | 40.2 | 35.5 |

\*Notes:
Cluster description: Cluster 1: Low vulnerable across all domains; Cluster 2: Primarily economic vulnerability; Cluster 3: Predominantly neighborhood-related vulnerability; Cluster 4: Economic and neighborhood vulnerability but favorable psychological conditions; Cluster 5: High vulnerability across all domains.
Abbreviations: COPD = Chronic Obstructive Pulmonary Disease; CKD = chronic kidney disease; BMI = Body Mass Index

Sex-specific differences in distribution across SDOH domains were observed. Female participants demonstrated a higher prevalence of economic disadvantage compared to their male counterparts (low income: 71.3% versus 59.2%; unemployment: 70.3% versus 60.2%) and psychological vulnerability indicators (living alone: 24.2% versus 19.0%; emotional distress: 53.3% versus 49.4%). Conversely, male participants exhibited greater social isolation (23.7% versus 14.3%) and reduced social support (29.0% versus 23.9%) compared to female participants. Neighborhood-level determinants demonstrated minimal sex difference, suggesting structural environmental factors operate more uniformly across sex groups.

### Identification of latent classes of SDOH

Measurement invariance testing favored the partially invariant model, which showed superior fit (Supplement Table S2). This supported the hypothesis that equivalent vulnerability constructs existed across sex groups while accounting for significant differences in distributional prevalence. We selected a 5-class approach because it appeared to fit the data best.

The distribution of SDOH indicators for each cluster is shown in Figure 1. Specifically, cluster 1 ("low vulnerable across all domains" n=972) showed the most favorable profile with no low-income participants and relatively low rates of psychological and neighborhood deleterious factors. Cluster 2 ("primarily economic vulnerability" n=1023) presented a unique pattern of lower household income (100%) and high unemployment (80.8%), but with relatively better performance in neighborhood factors and psychological stress levels. Cluster

**Figure 1.**
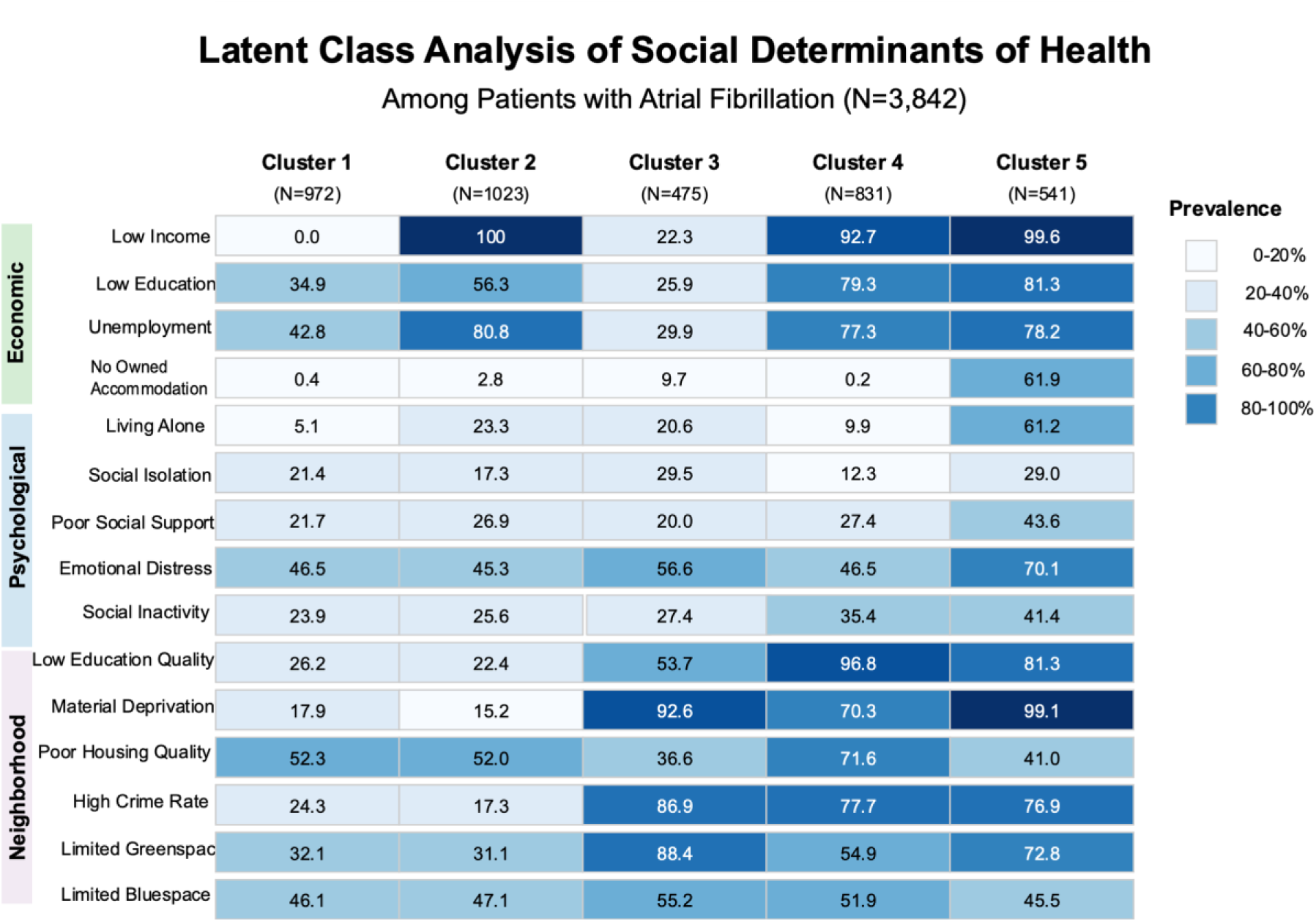
Distribution of latent class and conditional probability from single LCA

3 ("primarily neighborhood-related vulnerability" n=475) was distinguished by major neighborhood disadvantages including high material deprivation (92.6%) and higher crime rates (86.9%), despite moderate economic factors related vulnerabilities. Cluster 4 ("economic and neighborhood vulnerability but favorable psychological conditions" n=831) demonstrated high rates of poverty (92.7%) and poor neighborhood conditions, but relatively better psychological wellbeing with the lowest social isolation (12.3%). Finally, cluster 5 ("high vulnerability across all domains " n=541) emerged as the most disadvantaged group, characterized by higher rates of low income (99.6%), the highest rates of emotional distress (70.1%), social isolation (living alone 61.2%), and severe material deprivation (99.1%), despite having the highest rate of home ownership (61.9%).

Notable sex differences in vulnerability patterns were identified (Figure 2). Male participants demonstrated significantly higher representation in advantaged social profiles than female participants (Cluster 1: 28.3% vs 19.7%), suggesting greater likelihood of favorable SDOH. While economic-only vulnerability (Cluster 2) showed the opposite trend, with a higher percentage of females (31.4%) compared to males (24.1%). Of note, this class accounts for the largest proportion of females. Cluster 3 had the lowest representation in the total population, though slightly including more males (13.3%) than females (10.7%). Cluster 4 and Cluster 5 had relatively similar distributions between males and females, though females were slightly more represented in Cluster 4 (23.1% vs 20.8%) and males slightly more in Cluster 5 (15.1% vs 13.5%).

**Figure 2.**
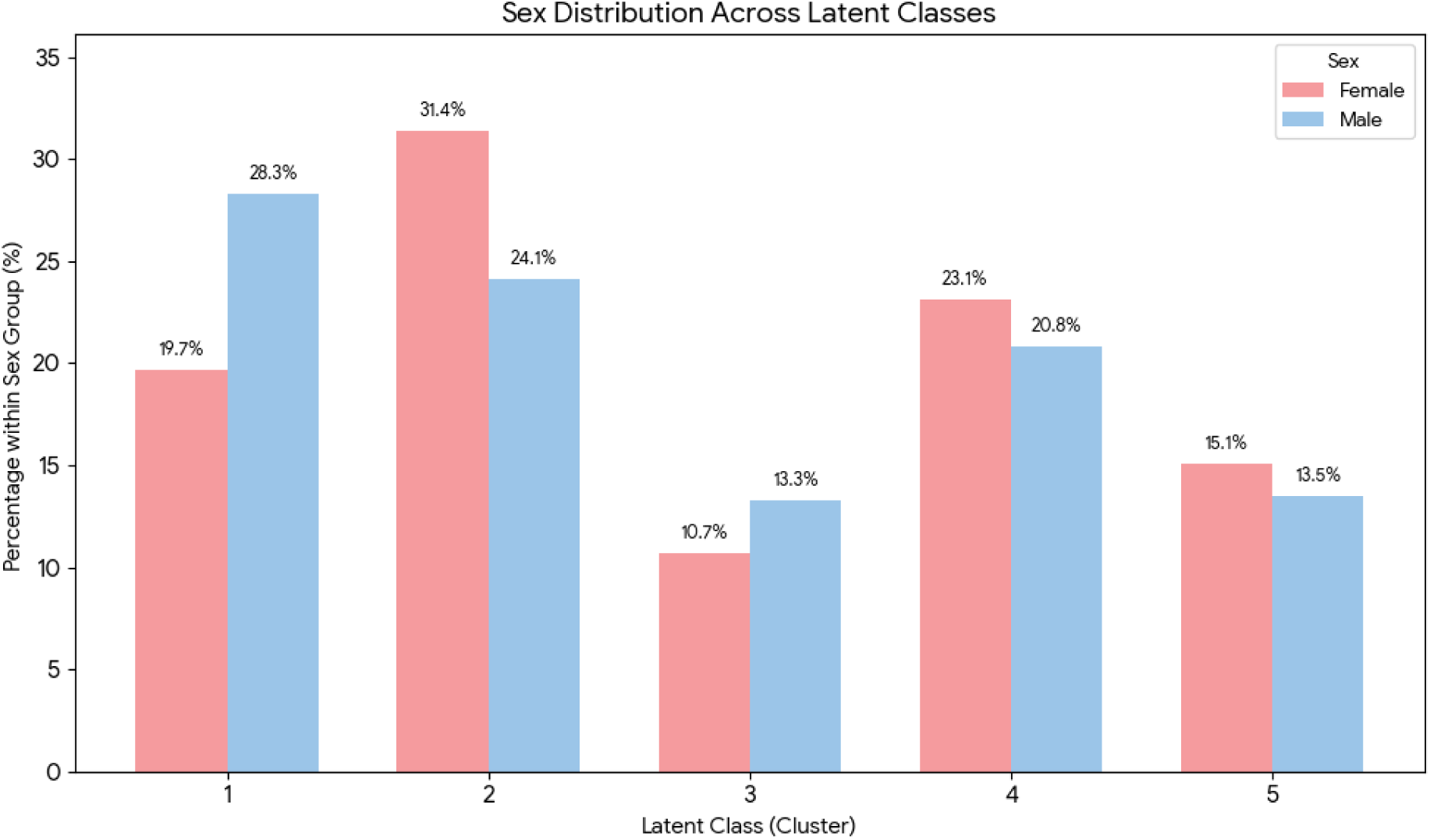
Sex distribution across latent class

### Association between clusters and outcomes

The association between the composite outcome and SDOH clusters were assessed. In a Cox proportional hazards analysis fully adjusted for clinical and behavioral variables, using Cluster 1 as comparison group, participants in Cluster 2 showed a 30% higher risk of the composite outcome (HR: 1.30, 95% CI: 1.07-1.58), while Cluster 4 demonstrated a 37% increased risk (HR: 1.37, 95% CI: 1.13-1.68). The most vulnerable group (Cluster 5) exhibited the strongest association, with more than doubled risk of MACE (HR: 2.05, 95% CI: 1.67-2.52). However, Cluster 3 showed no significant difference in outcome risk compared to Cluster 1 (HR: 1.04, 95% CI: 0.81-1.34).

Sex-stratified analysis revealed distinct SDOH-outcome association patterns (Figure 3). Female participants demonstrated a threshold effect pattern, with increased CV risk only for Cluster 5 (HR_female_=1.84, 95% CI: 1.22-2.76). In contrast, male participants exhibited a graduated risk pattern with significant elevation across vulnerability configurations: Cluster 2 (HR_male_=1.37, 95% CI: 1.09-1.71) and Cluster 4 (HR_male_=1.36, 95% CI: 1.08-1.71) demonstrated comparable risk magnitudes, while Cluster 5 yielded the highest risk (HR_male_=2.14, 95% CI: 1.69-2.72).

**Figure 3:**
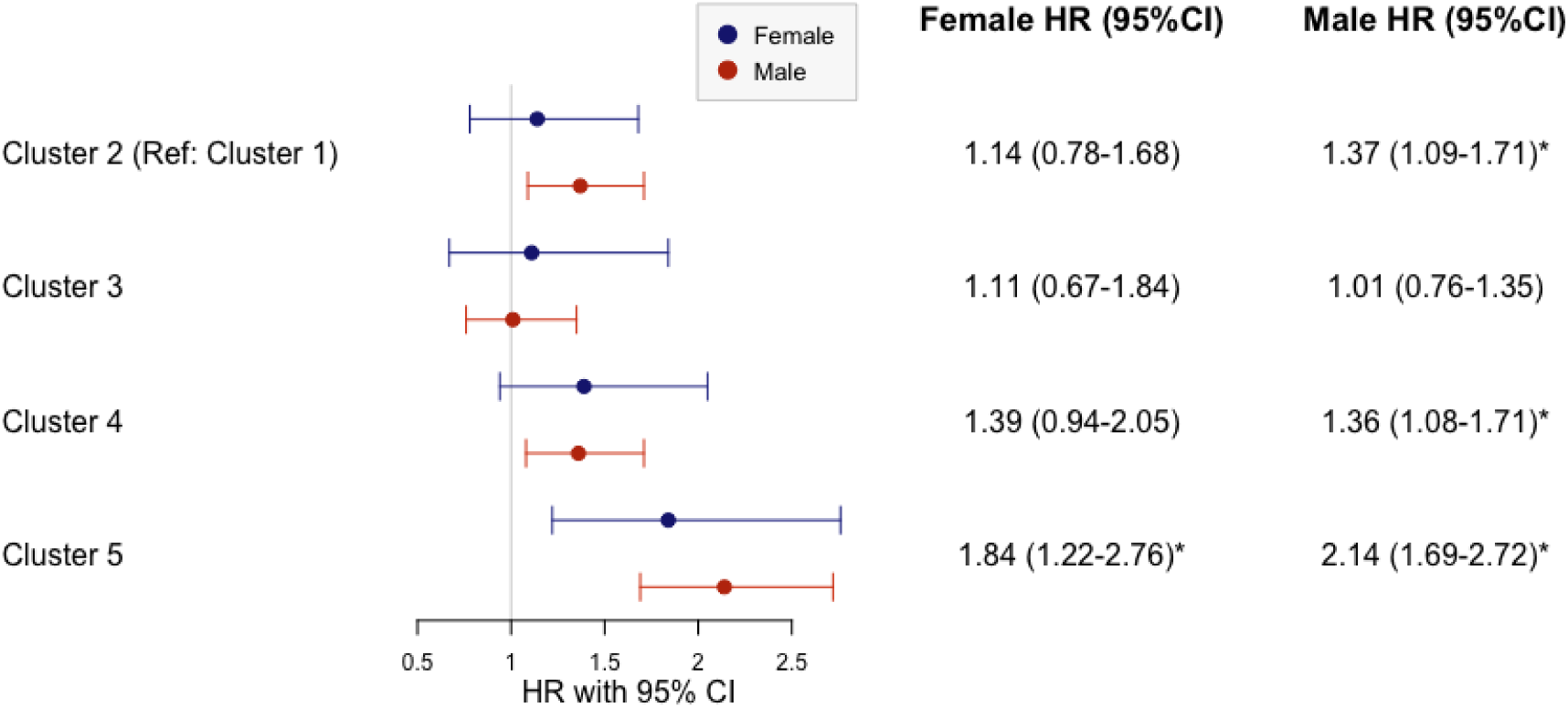
Forest plot for identified SDOH Cluster Associations with Composite Cardiovascular Outcome by Sex

Sensitivity analyses examining individual outcome components demonstrated similar risk hierarchies across clusters for CV mortality, non-fatal stroke, non-fatal myocardial infarction, and non-CV mortality, supporting the robustness of identified vulnerability-outcome associations across diverse CV endpoints.

## Discussion

This study of 3,842 individuals with AF applied multi-group LCA to identify SDOH clusters and their associations with MACE and all cause death. We identified five distinct patterns of social vulnerability through partially invariant multi-group models. The main findings are that economic factors constitute the predominant pathway linking social disadvantage to adverse CV outcomes, with clusters characterized by economic vulnerability consistently demonstrating elevated risk regardless of psychological or neighborhood factors. Furthermore, we observed that individuals with AF experienced similar SDOH clusters regardless of sex, however with different distributions across several clusters. To our knowledge, this is the first study to comprehensively identify patterns of SDOH using LCA specifically in individuals with AF, and to quantify their associations with CV outcomes. These findings underscore the complex interplay between SDOH and CV outcomes in AF participants.

Our findings demonstrated that economic disadvantage represents the predominant social factors associated with CV outcomes after adjusting demographic and clinical covariates. While various SDOH contribute to CV risk, our results point to the substantial impact of economic vulnerability. This is evidenced by the LCA where all classes with severe economic disadvantage (Clusters 2, 4, and 5) showed significantly increased composite outcome risk. These findings align with previous research highlighting the fundamental role of economic status in CV health (30,31). For instance, findings from the Prospective Urban Rural Epidemiologic (PURE) study, a large international prospective cohort study, showed that participants with low income and low education are up to two times more likely to experience MACE and 2 to 3 fold more likely to experience CVD-associated mortality, relative to those with favorable socioeconomic profiles (32). The impact of economic vulnerability on CV outcomes likely operates through multiple pathways. First, economic constraints may affect medication adherence, particularly for expensive anticoagulants and antiarrhythmics that are crucial for stroke prevention in individuals with AF (9). Second, despite universal healthcare coverage in the UK, individuals’ economic barriers still affect healthcare utilization through indirect mechanisms such as transportation costs to appointments, inability to take time off from hourly-paid work, digital exclusion limiting access to remote monitoring services, and challenges navigating complex healthcare systems without adequate support (33). Third, economic hardship may lead to chronic stress and adoption of adverse health behaviors, such as smoking and poor dietary choices, which were more prevalent in our economically vulnerable groups (30). Additionally, economic status is associated with education, health literacy and self-management capabilities, which are essential for successful AF care (9,34,35).

Interestingly, our finding that isolated neighborhood disadvantage (Cluster 3) was not linked to cardiovascular outcomes differs from what has been reported in some established social epidemiology studies. (31). For instance, Diez Roux et al. found that residents of disadvantaged neighborhoods had a higher risk of coronary heart disease, even after adjusting for personal income, education, and occupation(36), and Abdel-Qadir et al. demonstrated associations between neighborhood deprivation and CV mortality (33). Several factors might explain the apparent discrepancy between our findings and previous research. First, participants with better economic resources might be able to overcome neighborhood limitations through better access to transportation and healthcare services outside their immediate area (37). Second, they may have greater capacity to modify their home environment or more likely to engage in healthier behaviors to mitigate neighborhood disadvantages (16). Third, the protective effects of economic stability might buffer against the negative impacts of poor neighborhood conditions (38,39). These findings may suggest that while neighborhood factors contribute to CV risk, their impact might be less direct or more modifiable than individual-level economic factors among individuals with AF.

Multi-group LCA revealed vulnerability constructs maintain identical structural characteristics across sexes while accommodating significant distributional prevalence differences. This methodological achievement enables meaningful cross-group comparisons while ensuring construct validity, essential prerequisites for evidence-based intervention development and health policy formulation. The finding that sex-based heterogeneity manifests primarily through distributional variations provides important insights regarding SDOH operational mechanisms. It aligns with previous view that different patterns exist in lifestyle, health beliefs, and behaviors among males and females (40). These findings underscore the importance of considering sex-specific factors when designing health interventions and support systems (41). For instance, interventions aimed at enhancing resilience and health managing might need to be tailored differently for males and females to be effective.

### Strength and Limitations

This study is novel as it applies an intersectional and multidimensional-based approach to explore the interaction between SDOH, sex and CV outcomes in a large sample of AF participants. This study highlights the ways in which SDOH may exert detrimental effects on outcomes that differ by sex.

However, our findings should be interpreted with caution considering several limitations. First, the cross-sectional design of our study only provides a snapshot of social circumstances at one point in time, limiting our ability to understand how long-term exposure to social disadvantage affects CV risk. Second, the direction of causality cannot be definitively established, as declining health might lead to changes in social and economic circumstances (such as unemployment or social isolation)(42), potentially creating bidirectional relationships between SDOH and health outcomes. Third, while we adjusted for multiple clinical factors, unmeasured confounding may still exist. For example, health literacy and health insurance usage were not directly measured but could be associated with both SDOH patterns and CV outcomes(34). In addition, as our study was conducted in UK with a universally inclusive healthcare system, our findings may not be generalizable to individuals with AF in different healthcare systems or social contexts. Furthermore UK Biobank participants are known to be generally healthier and more educated than the UK population, with implicit selection cohort biases (43). Future research would be enhanced by longitudinal assessment of SDOH patterns, more detailed measurement of social circumstances, and validation in diverse populations.

## Conclusion

In conclusion, we identified five distinct SDOH clusters in individuals with AF using multi group LCA. The identification of economic vulnerability as the predominant factor associated with adverse CV outcomes provides clear targeting opportunities for healthcare system interventions designed to address modifiable SDOH in AF population. These findings position economic barrier mitigation as the leading strategy for reducing cardiovascular risk in individuals with AF, underscoring the need for evidence-based interventions and optimized healthcare delivery.

## Ethical approval

Study data were obtained from the UK Biobank data resources. The UK Biobank study received approval from the Northwest Multi-Centre Research Ethics Committee (11/NW/0382), https://www.ukbiobank.ac.uk/learn-more-about-uk-biobank/about-us/ethics). Participants provided informed consent at baseline. Our analyses were conducted under the UK Biobank application number 45551.

## Author Contribution

Data curation was done by YZ. Conceptualization, funding acquisition, investigation, methodology, project administration, resources, software, supervision, validation, and visualization were conceived by YZ, JH, LP. YZ and JH did the formal analysis and wrote the initial draft of the manuscript. All authors contributed substantially to the discussion, reviewing and editing, and approved the final manuscript.

## Data availability

Data is available on request.

